# Mechanical Ventilator Milano (MVM): A Novel Mechanical Ventilator Designed for Mass Scale Production in Response to the COVID-19 Pandemics

**DOI:** 10.1101/2020.03.24.20042234

**Authors:** C. Galbiati, W. Bonivento, M. Caravati, S. De Cecco, T. Dinon, G. Fiorillo, D. Franco, F. Gabriele, C. L. Kendziora, I. Kochanek, An. Ianni, A. McDonald, L. Molinari Tosatti, M. Malosio, D. Minuzzo, S. H. Pordes, A. Prini, A. Razeto, M. Razeti, M. Rescigno, D. Sablone, E. Scapparone, G. Testera, R. Tartaglia, H. Wang, A. Zardoni

## Abstract

We present here the design of the Mechanical Ventilator Milano (MVM), a novel mechanical ventilator designed for mass scale production in response to the COVID-19 pandemics, to compensate for the dramatic shortage of such ventilators in many countries. This ventilator is an electro-mechanical equivalent of the old, reliable Manley Ventilator. Our design is optimized to permit large sale production in short time and at a limited cost, relying on off-the-shelf components, readily available worldwide from hardware suppliers. Operation of the MVM requires only a source of compressed oxygen (or compressed medical air) and electrical power. The MVM control and monitoring unit can be connected and networked via WiFi so that no additional electrical connections are necessary other than the connection to the electrical power.

At this stage the MVM is not a certified medical device. Construction of the first prototypes is starting with a team of engineers, scientists and computing experts. The purpose of this paper is to disseminate the conceptual design of the MVM broadly and to solicit feed-back from the scientific and medical community to speed the process of review, improvement and possible implementation.

## 1. INTRODUCTION

The current COVID-19 pandemic [1] is causing a rapidly increasing number of SARS-CoV-2 [2]pathologies all across the world. A common outcome of SARS-CoV-2 is severe pneumonia [3]. Effective medical intervention in this pathology involves the use of mechanical ventilation [3–5]. In order to meet the needs of the increasingly large number of patients in the COVID-19 pandemic, the rapid deployment of a large number of new mechanical ventilators [6] will be required.

These devices must be capable of being mass produced rapidly and at large scale.

There are substantially two different modalities of mechanical ventilation [7]:

### Positive end-expiratory pressure (PEEP)

a ventilation mode where *“the pressure in the lungs (alveolar pressure)”* at the end of the expiratory cycle is maintained *“above atmospheric pressure (the pressure outside of the body)”* [8]. *“The two types of PEEP are extrinsic PEEP (PEEP applied by a ventilator) and intrinsic PEEP (PEEP caused by an incomplete exhalation). Pressure that is applied or increased during an inspiration is termed pressure support”* [8];

### Pressure Support Ventilation (PSV)

*“a spontaneous mode of ventilation. The patient initiates each breath and the ventilator delivers support with the preset pressure value. With support from the ventilator, the patient also regulates his own respiratory rate and tidal volume”* [9]. See also Ref. [10] for additional information.

The imperative need to treat a large number of patients affected by SARS-CoV-2 has motivated the consideration of a reliable, fail-safe, and easy to operate mechanical ventilator that can be produced quickly on a massive scale. We introduce a novel mechanical ventilator (Mechanical Ventilator Milano, MVM), inspired by the Manley ventilator [11]. In 1961, Roger Manley *“suggested the possibility of using the pressure of the gases from the anaesthetic machine as the motive power for a simple apparatus to ventilate the lungs of the patients in the operating theatre”* [12]. We have tried to design the MVM with the same principle of simplicity in mind. The main difference is the use of electrically-driven pneumatic valves instead of mechanical switches to ease large scale production. The MVM does not include a system to measure the tidal volume provided to the patient with each breathing cycle, but rather a system to mea-sure the peak and average expiratory flow rate. The MVM adheres as much as possible to the guidelines for “Rapidly Manufactured Ventilator System (RMVS)” [13] recently released by the UK Medicine & Healthcare products Regulatory Agency.

The MVM delivers to patients PEEP mechanical ventilation, reaching a pressure in the range 20 mbarg to 30 mbarg (1 mbarg is the atmospheric pressure plus a pressure given by a H_2_O column of 1 cm of height) at the end of inspiration and an end-expiratory pressure of approximately 5 mbarg. The MVM can operate in either an independent or in assisted mode, described later in Sec. 3.

The main characteristics of the MVM are:

### Simplicity of Operation

the MVM connects directly to a line of pressurized medical oxygen (or medical air), and relies on simple regulation of the upstream flow to deliver oxygen to the patient at a pressure in the range 20 mbarg to 30 mbarg/cm H_2_O. Pressure regulation of the end-expiratory cycle is achieved by discharging the expiratory flow in a simple vent trap, which sets the desired minimum expiratory pressure precisely at 5 mbarg/cm H_2_O. Another vent trap connected to the line delivering oxygen to the patient ensures that the maximum pressure delivered does not exceed 30 mbarg. The construction of both vent traps with transparent plastic containers permits to doctors and nurses a direct visual monitoring of the inspiratory pressure and of the expiratory flow;

### Small Number of Components

the MVM consist of a medical care flowmeter [14] in use for direct regulation of the maximum flow rate, a oxygen-therapy humidifier [14], a non-vented non invasive ventilation (NIV) mask [15], two electrically-controlled, VDC-actuated pneumatic valves, a pressure sensor, a control system directly connected to the pneumatic valves and to the control sensor, and a backup VDC battery connected to the control system;

### Ease of Procurement

the parts required for the construction of the MVM have been selected by investigating current inventories available worldwide at major resellers of hardware suppliers. The parts selected are also characterized by their ease of mass-scale manufacturing;

### Ease of Construction

assembly of the parts into a complete MVM can be completed with a set of limited and clear instructions. The process for loading the software into the controller is simple. The controller software is open source and available for customizations by end users;

### Cost Containment

the total cost of components is a few hundreds of € ‘s.

### Ease of Deployment

the only required facilities are a line of pressurized oxygen and standard AC electric power (either 220 V or 110 V). The MVM can be deployed wherever a source of pressurized oxygen can be made available. It is suitable for home care and for use in ambulances. It is convenient to deploy in care clinics with centralized oxygen supply systems, such as COVID hospitals or COVID-care areas in general hospitals;

### Customizability

the MVM can operate in different configurations (independent and patient assisted) as described in Section 3. The operating parameters can be tuned by the operator with a simple user interface;

### Scalability

the MVM control system is based an a WiFi micro-controller which allows to monitor and tune the parameters of multiple units with a single work station;

### Reliability

all the components in this units are commercial and readily available. The reliability of MVM has to be carefully studied based on the specifications of the components.

The system is designed to be simple, cheap and easily reparable, just by replacing the non-functional parts.

## 2. DESIGN AND COMPONENTS

The conceptual design and P&ID of the MVM are shown in Fig.1. The main components are described following the order of the oxygen flow:

**FIG. 1.**
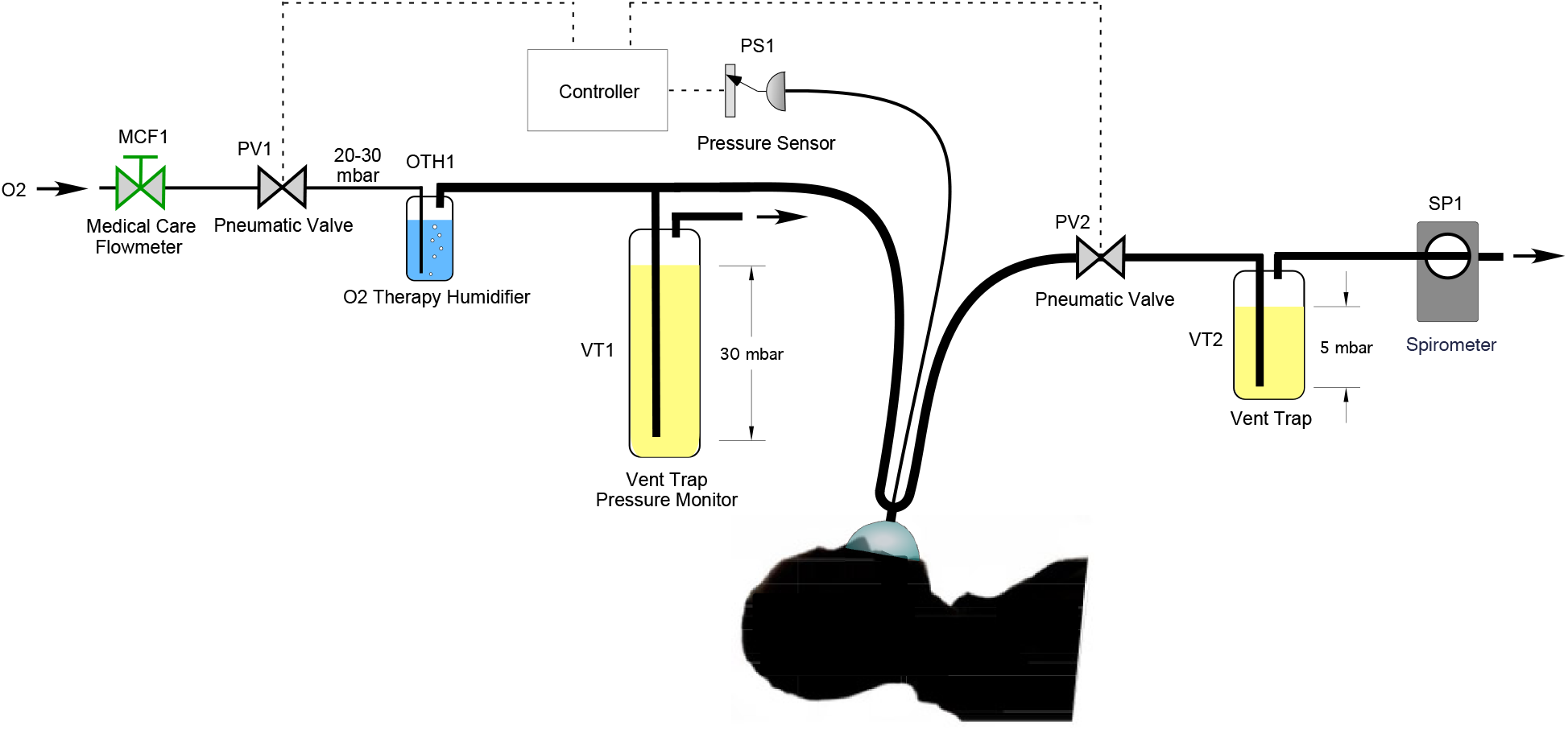
Conceptual design and P&ID of MVM. Note: 1 mbar corresponds to the pressure of a H_2_O columns of 1 cm height.

**FIG. 2.**
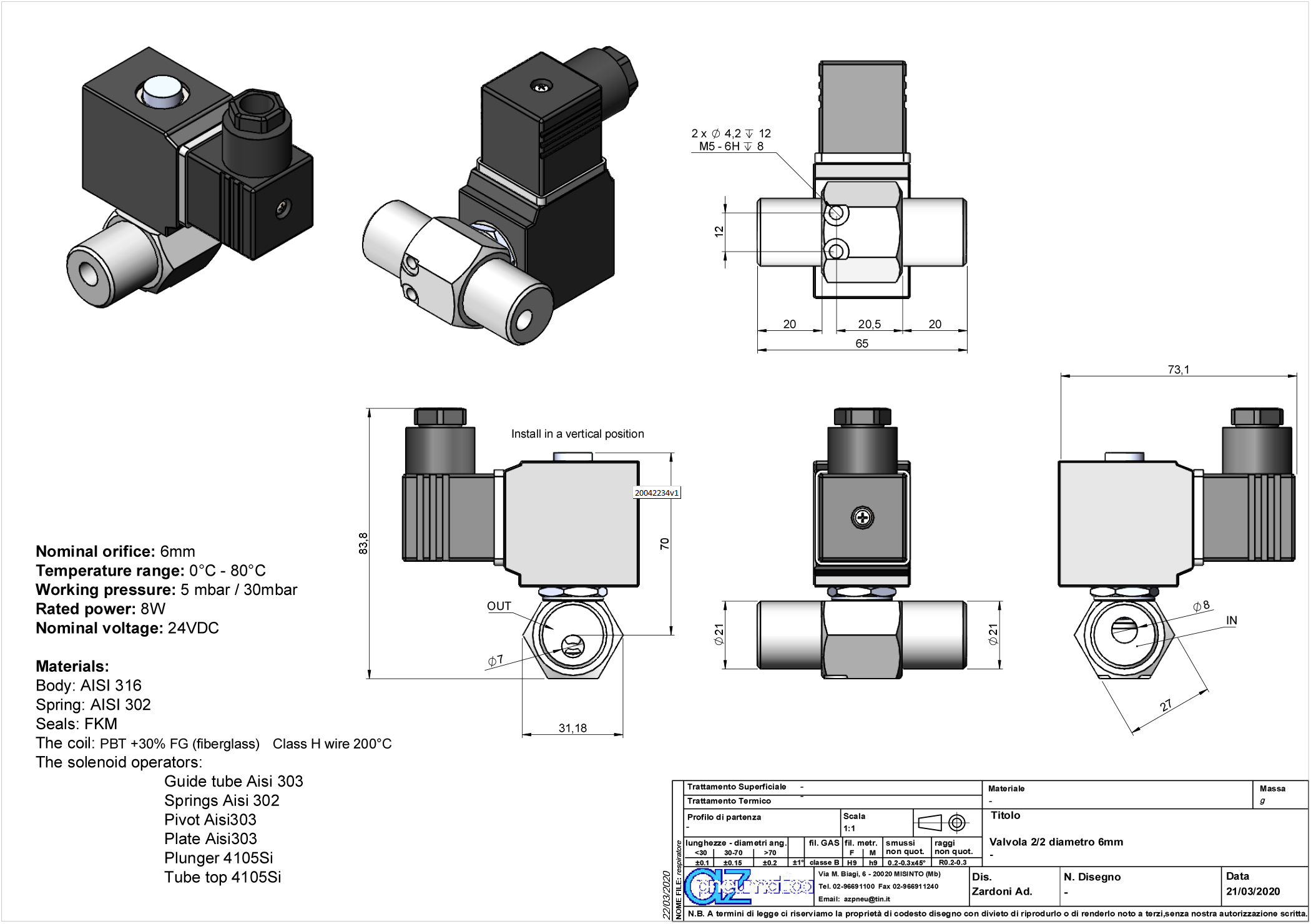
Data sheet [17] of a possible valve in use for the MVM. Two valves per unit are required. The termination of the valves are compliant with the standard for 22 mm cones of ISO-5356-1:2015 [16].

**FIG. 3.**
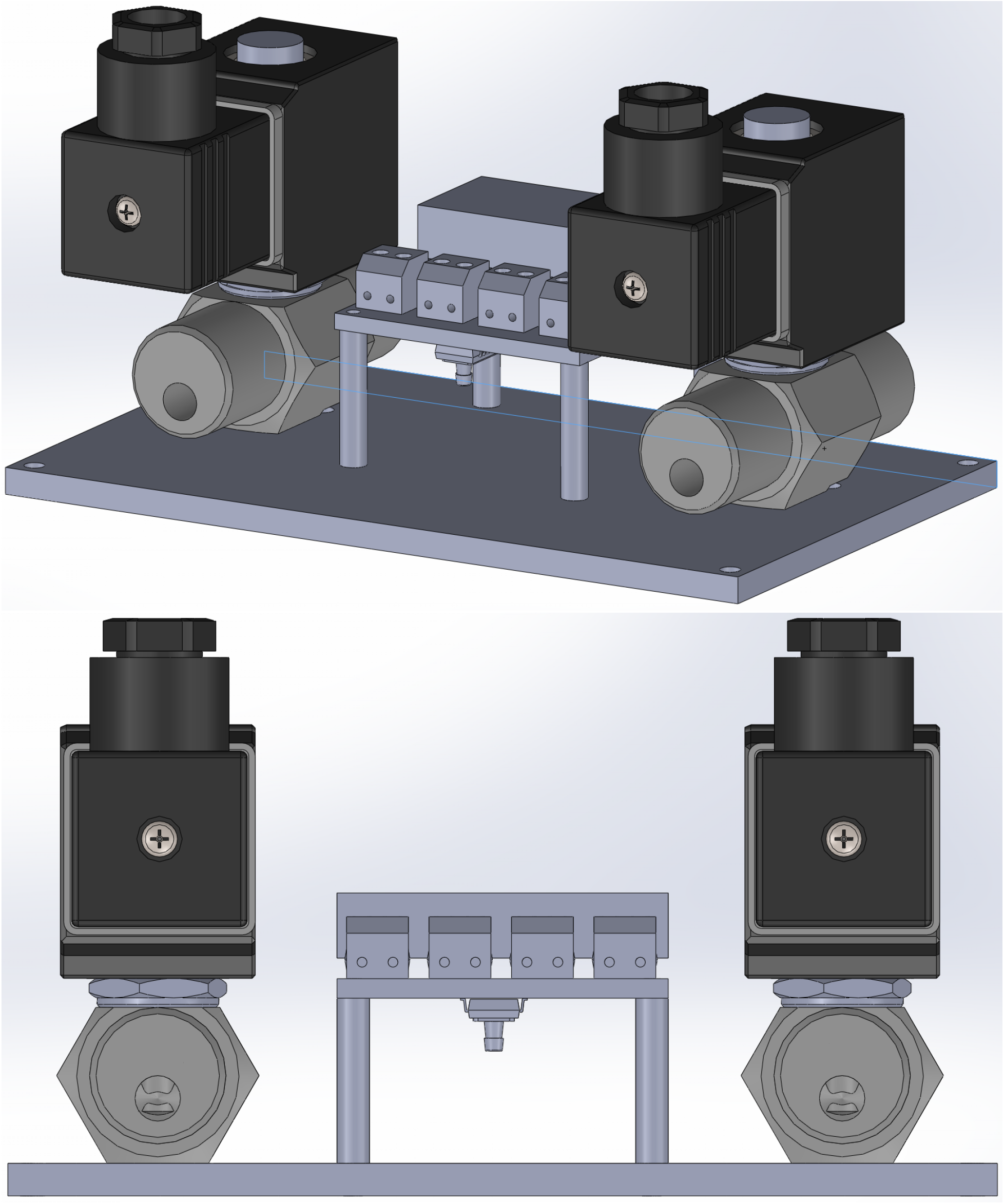
MVM control unit, in its 3D view (top) and lateral view (bottom). To give a scale,the base plate is 152 mm by 170mm

### Connection to oxygen supply

at the left end side, the MVM is connected to a pressurized oxygen line.

### Medical care flowmeter MCF1

The oxygen flow is controlled by medical care flowmeter MCF1, which restricts the flow to a maximum value. In case of the need to administer a mix of oxygen and air, MCF1 can be an oxygen/air mixing medical care flowmeter.

### Oxygen delivery pneumatic valve PV1

The incoming oxygen flow is controlled by pneumatic valve PV1. We have verified that an orifice with diameter of 6 mm is sufficient to guarantee that a proper respiratory *minute volume* is delivered to the patient;

### Humidifier OTH1

the incoming oxygen flow is humidified before reaching the patient by entering the oxygen therapy humidifier OTH1;

### Vent trap VT1

vent trap VT1 is filled with a vertical column of oil, which raises the minimum vent pressure to a value of approximately 30 mbarg. The value of the minimum vent pressure can be easily adjusted in the neighborhood of 30 mbarg by varying the oil level or by adjusting the vertical position of the tube reaching below the oil surface. The vent line connects directly to the atmosphere, guaranteeing that the pressure differential delivered to the patient does not exceed the preset minimum vent pressure of VT1;

### Nasal or oronasal mask

from OTH1, the incoming humidified oxygen flow is delivered to the patient outfitted with a non-vented NIV mask.

### Breathing system

the NIV mask is connected to a breathing system, supporting the attachment of two plastic tubes of typical size 22 mm connecting respectively to pneumatic valves PV1 and PV2, and to a smaller plastic tube leading to pressure sensor PS1. The standard for connection of the breathing system is the 22 mm cone and socket combination defined in the publication *“ISO 5356-1:2015 - Anaes-thetic and respiratory equipment — Conical connectors — Part 1: Cones and sockets”* [16].

### Oxygen line

the oxygen line from OTH1 to the patient is branched into a line connected to vent trap VT1;

### Expiration pneumatic valve PV2

PV2 controls the expiratory flow. As for PV1, a valve orifice with diameter of 6 mm is sufficient to guarantee the flow corresponding to the expiration of a proper respiratory minute volume;

### Vent trap VT2

vent trap VT2 is filled with a vertical column of liquid, which raises the minimum vent pressure to the value of 5 mbarg;

### Spirometer SP1

a portable digital spirometer can be connected to the vent line of vent trap VT2 to monitor the peak and average expiratory flow rate. We are also investigating the possibility to integrate the spirometer in the main control unit, shifting its installation point so that it is directly connected to the downstream port of PV2;

### Note on VT1 and VT2

both those liquid based vent traps act as adjustable pressure relief valves where the pressure level can be monitored. They could be replaced by mechanical pressure relief valves set at fixed values of 30 and 5 mbarg respectively. They are very compact objects available on the market, the only drawback being the pressure in the ventilation line cannot be monitored.

## 3. CONTROL SYSTEM AND OPERATION

The control system performs supervision and actuation of the two valves based on the programmed respiratory cycle.

The respiratory cycle is operated by actuating pneumatic valves PV1 and PV2. During the inspiratory phase, PV2 is closed and PV1 is open. At the end of the inspiratory cycle, the pressure reaches the design range of 20 mbarg to 30 mbarg. At the end of the inspiratory cycle, PV1 is closed and PV2 is opened simultaneously, allowing the discharge of the lungs pressure. The end-expiratory pressure is set by the height of the liquid column in vent trap VT1.

Pressure sensor PS1 reads directly the pressure at the mask of the patient and permits the control system to verify and ensure that the pressure is always within the operating range by operating the relative valves: 20 mbarg to 30 mbarg for the inspiration phase and 5 mbarg for the end-expiratory phase.

The main controller runs on a (`Arduino`-compatible) micro-controller board based on a 32 bit micro-controller with WiFi/Bluetooth connectivity. All boards of this type have a small form factor and integrates all I/O function required for this project.

A daughter board interfaces to the controller and provides four opto-coupled switches to operate the electrically-controlled pneumatic valves. The daughter board is provided with 24 V VDC supply and includes the low voltage regulator to supply the central unit.

The differential pressure gauges are based on a 60 mbar differential pressure sensor with a resolution better than 0.2 mbar.

The WiFi connectivity of the board the unit offers the option for central monitoring and configuration. A buzzer and a high luminosity LED are available to signal the operators in case of alarms.

The software operating the breathing cycles will be defined in conjunction with medical doctors involved in the development of the units, and will allow operation in independent or assisted configuration. In its independent configuration the unit will operate the valves at fixed cycles as configured by the operator. In its assisted configuration, the cycles will be defined by a feedback system that will detect the incipient movement of the thoracic cage by monitoring with high precision the pressure at the mask through pressure sensor PS1. Upon detecting the incipient movement of the thoracic cage, the MVM will trigger the assisted PEEP breathing cycle movement.

The MVM is equipped with an industrial power supply unit capable for at least 50 W and battery backup operated in fail-safe mode. Under normal circumstances the power supply will feed the controller and keep the battery charged. In case of power failure the battery will automatically intervene and provide the current to operate the system for an interval not in excess of 30 min. The power supply is hosted in a separate enclosure to provide isolation between the oxygen lines and possible spark sources. The power supply unit will include two 12 V, 1.2 A h batteries.

## 4. STATUS OF DEVELOPMENT

We thank Prof. A. Pesenti of Universitá degli Studi di Milano, Dr. R. Guerra of World Health Organization, and R. Pordes for discussion and comments. The first functional tests were performed on Friday, March 20, 2020, utilizing mechanical ventilators in lieu of vent traps to set the upstream and downstream pressures and a silicone bag test lung. Videos from the demonstration session are available at https://tinyurl.com/uurapy3. Engineering towards the first prototype units started on Saturday, March 21, 2020, and construction of the first prototype units on Monday, March 23, 2020. A preliminary list of candidate components is given in Appendix A.

## Data Availability

there are no data

## Appendix A Candidate Components

### Micro-controller board

A practical example is the `Huzzah ESP32` [18] from `Adafruit;`

### Medical care flowmeter

Practical examples are the EasyFlow^©^ or Qmed^©^ medical care flowmeters shown at Ref. [14];

### Mixing Medical Care Flowmeter

the EasyMIX^©^ unit shown in Ref. [19];

### Non-vented NIV mask

One practical example is the NIV oronasal mask NovaStar^©^ TS AAV without exhalation ports, catalog number MP04701-04 from `Dräger`, page 64 of Ref. [15];

### Breathing system

One practical example of a breathing system is the breathing circuit VentStar^©^, disposable, catalog number MP00338 from `Dräger`, page 11 of Ref. [15];

### Electrical power supply

Convenient examples are the integrated switching power supply TXL 060-24S from `TRACO POWER` [20] and the battery code 537-5444 from `RS PRO` [21].

